## Supplemental Figures and Tables for "Micronutrient-deficient diets and possible environmental enteric dysfunction in Buruli ulcer endemic communities in Ghana: lower dietary diversity and reduced serum zinc and vitamin C implicate micronutrient status a possible susceptibility factor"

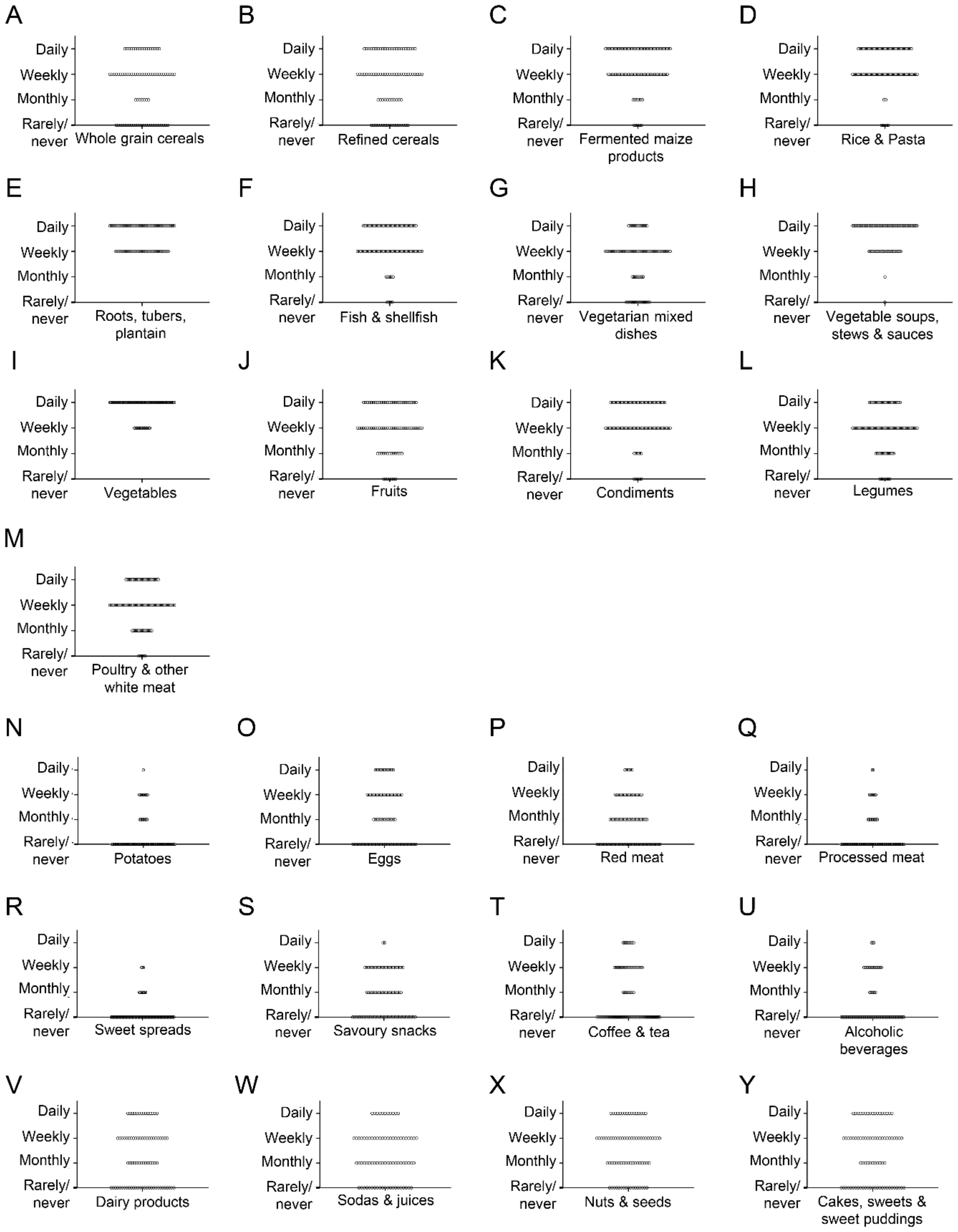


**S1 Figure. The most and least frequently consumed food groups by the study participants.** A food frequency questionnaire was put to all 80 Cohort 1 participants (40 BU cases, 40 controls), covering a total of 105 foods from 25 food groups.  There are 4 possible answers for frequency of consumption for each food, namely daily, weekly, monthly, and rarely/never. This figure visualises the distribution of the most frequent consumption of one or more foods in that food group in the past month amongst the participants. The red line represents the frequency of consumption, and its length is proportional to the number of participants with that maximal frequency. Data from the food groups showed that at least 60% of the participants ate foods from these food groups at least weekly, where similar numbers consumed foods monthly, rarely or never vs. weekly or daily (N-Q), and those where at least 60% of the participants ate them monthly, rarely or never (R-Y).

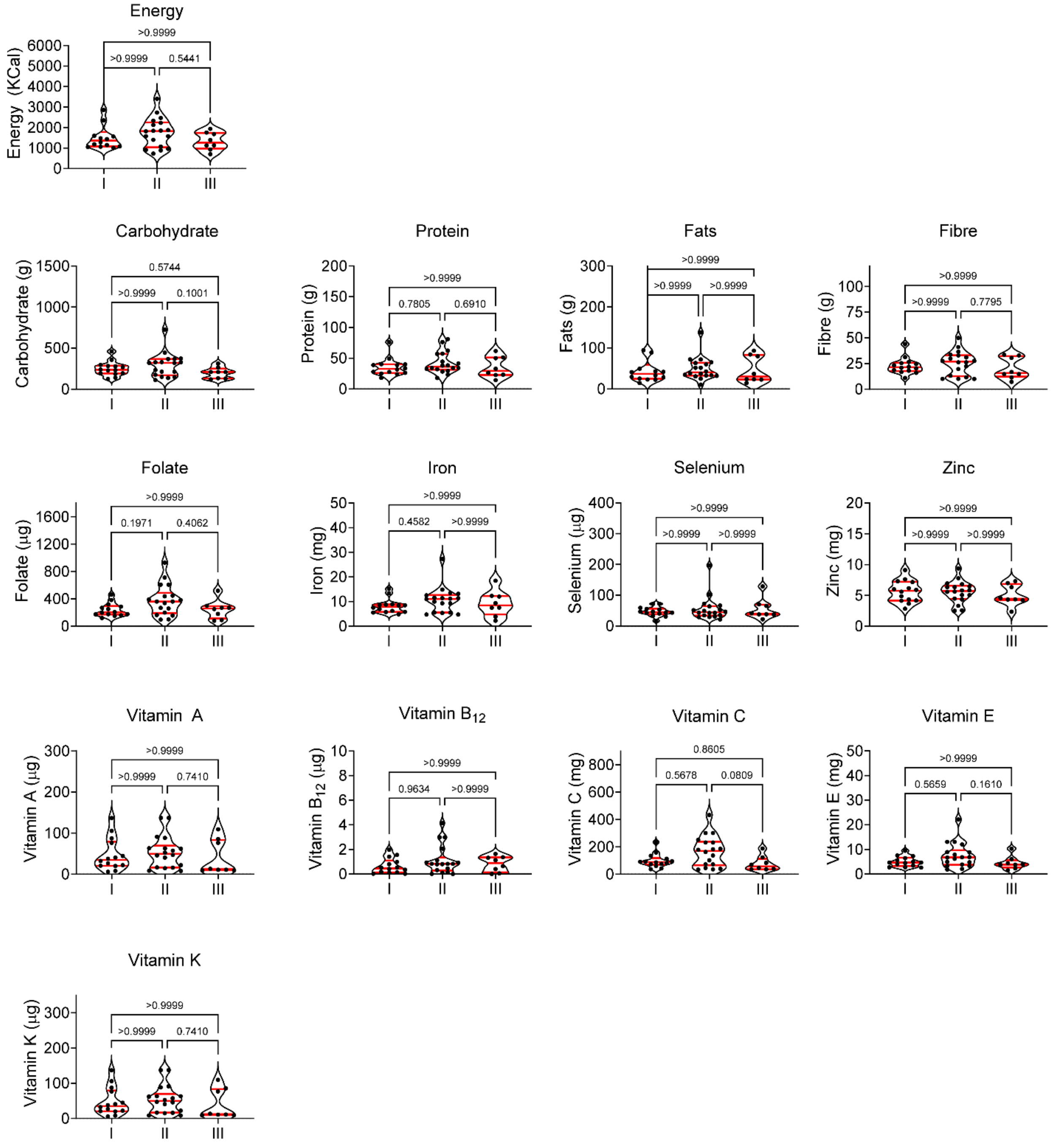


**S2 Figure. No difference in nutrient intake between BU cases with different category of lesion.** Median, interquartile range comparison of the category of lesions to nutrient intake was analysed using the Kruskal-Wallis test and Dunn post-hoc test for non-parametric data for comparison between category I versus II lesions, category I versus III lesions, category II versus III lesions individually.

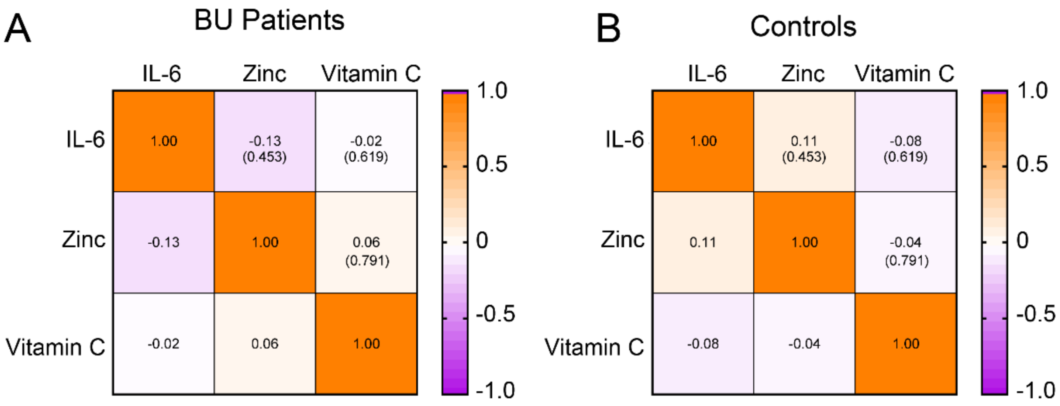


**S3 Figure. No correlation between nutrition and inflammation markers in BU cases and controls.** Serum concentrations of vitamin C, zinc and IL-6 were analysed using Spearman's correlation, and are presented as a heatmap. Spearman's correlation coefficients are given along with P-value (in brackets).

**S1 Table. List of foods used in the FFQ administration.**

| Food group | Food items |
|---|---|
| Whole grain cereals | Oats porridge, maize porridge, millet porridge, wheat porridge, whole meal bread/roll, Tuo zafi (T.Z.) |
| Refined cereals | Sugar coated cereals, cornflakes, white bread/roll, pancake, flour chips/achumo, kaafa |
| Fermented maize products | Kenkey, Banku, Akple |
| Rice and pasta | White rice cooked, local rice cooked, noodles, macaroni |
| Roots, tubers and plantain | Yam, cassava or cocoyam, plantain, fufu, kokonte, gari |
| Potatoes | Boiled instant potatoes/oven baked potatoes, fried potatoes chips, roast potatoes |
| Nuts and seeds | Cashew nuts, groundnut/pea nuts, almond/hazelnut, pistachio, coconut, sultana/raisins/currants |
| Legumes | Kidney beans, soya beans, black eyed beans, baked beans, chicks peas, runner/green beans, ground nut soup |
| Dairy products | Milk, full fat yoghurt, full fat cheese, low fat cheese, butter/margarine, cocoa, horlicks, milkshake/fula |
| Eggs | Egg boiled/poached, egg fried/omelette |
| Red meat | Beef/lamb/pork, bush meat, rabbit |
| Poultry/other white meat | Chicken/other poultry, snail |
| Processed meat | Burger, bacon, ham, sausage/luncheon meat/corned beef |
| Fish *and shellfish* | Fish, fish fingers, fish fried in butter, crabs, prawn, octopus |
| Vegetarian mixed dishes | Jollof rice, fried rice, waakye, soya bean curd/tofu, beans stew |
| Vegetable soups, stews, sauces | Tomato stew, melon seed and spinach stew, palmnut soup, okro soup/stew, light soup, kontomire (cocoyam leaves) |
| Vegetables | Sweet corn, okro/nkruma, garden egg/aubergine, avocado, garden peas/peas, mushroom, broccoli/cauliflower, carrot, cabbage, brussels sprouts, ayoyo, spring onions, onions, tomatoes, tin-tomato, sweet peppers, lettuce, cucumber, coleslaw, beetroot, green pepper, Ghanaian green pepper (Kpakposhitor), |
| Fruits | Banana, pears, pomegranates, mangoes, pineapple, grapefruits, orange/satsuma, grapes, melon, apple, peach/plum/nectarine, strawberries/cherries, pawpaw |
| Sweet spreads | Jam, marmalade, honey, peanut butter |
| Cakes and sweets, and sweet puddings | Sugar added to cereals, tea, coffee, cakes/scones/doughnuts, ice cream/frozen desserts, sweets biscuits, chocolate coated sweet biscuits, chocolate/chocolate bar, custard, rice pudding, sweets, toffees, mints |
| Savoury snacks | Crisp/pocket snacks, fried plantain chips, salty biscuits, regular popcorn, buttered popcorn |
| Condiments | Hot pepper sauce, tomato ketchup, salad pepper (powdered) |
| Alcoholic beverages | Beer/larger/cider, port/sherry/liqueur, spirits, pito/solom, palmwine (alcohol) |
| Sodas and juices | Real fruit juice, fruit squash, fizzy soft drink, low calorie fizzy drink, bissap (sobolo), liha (sprout maize), palmwine (fresh) |
| Coffee and tea | Tea, Coffee |

**S2 Table. Food groups used in the Dietary Diversity Score, generated from 24-hour recall data.**

| | | |
|---|---|---|
| cereals | roots & tubers | green leafy vegetables |
| other vegetables | fruits | meat |
| fish & seafood | eggs | pulses & nuts |
| milk & milk products | oils & fats | sugar |
| condiments | drinks & beverages | ‘other’ |

**S3 Table. Socioeconomic status of Cohort 1 study participants.**

| | **Cohort 1** | | |
|---|---|---|---|
| **Variables** | **BU cases**<br>n=40 | **Controls**<br>n=40 | **P-value** |
| **Education** | | | 0.236[a] |
| *Primary* | 24 (58.5%) | 18 (45%) | |
| *Secondary* | 8 (19.5%) | 16 (40%) | |
| *Tertiary* | 0 | 2 (5%) | |
| *None* | 9 (22%) | 4 (10%) | |
| **Marital Status** | | | 0.769[a] |
| Schooling | 20 (50%) | 22 (55%) | |
| *Single* | 6 (15%) | 6 (15%) | |
| *Married* | 13 (33%) | 12 (30%) | |
| *Divorced* | 1 (3%) | 0 | |
| **Household monthly income** | | | 0.478[a] |
| *<GH₵ 50* | 6 (15%) | 4 (10%) | |
| *GH₵ 50-200* | 10 (25%) | 15 (38%) | |
| *GH₵201-800* | 23 (58%) | 21(53%) | |
| *>GH₵800* | 0 | 0 | |
| **Occupation** | | | |
| *Formal* | 0 | 0 | 0.236[a] |
| *Informal* | 15 (38%) | 17 (43%) | |
| *Attending School* | 20 (50%) | 22 (55%) | |
| *Unemployed* | 5 (13%) | 1 (3%) | |

[a]; T $\chi^2$ test. p-value < 0.05 was statistically significant.

**S4 Table. Mean percentage of macronutrients in the diet of Cohort 1 participants.**

| Nutrients | Mean nutrient intake (%) | | |
|---|---|---|---|
| | **All**<br>N=80 | **BU Cases**<br>N=40 | **Controls**<br>N=40 |
| Energy (Kcal) | 1629 (100%) | 1594 (100%) | 1664 (100%) |
| Carbohydrate (g) | 292 (64.4%) | 261.9 (64.4%) | 322.7 (67.9%) |
| Protein (g) | 42.2 (9.6 %) | 38.84 (9.6%) | 45.63 (9.6%) |
| Fats (g) | 23.5 (24.0%) | 46.99 (26.0%) | 47.52 (22.5%) |

Unit conversions: 1g of carbohydrate is equivalent to 4 Kcal, 1g of protein is equivalent to 4 Kcal and 1g of fats is equivalent to 9 Kcal, which were then used to calculate percentages.

**S5 Table. Comparison of energy and nutrient intake of Cohort 1 study participants between sexes.**

| Nutrients | Sex | BU Cases<br>Male N=19; Female N=21<br>Median (range) | P-value | Controls<br>Male N=22; Female N=18<br>Median (range) | P-value |
|---|---|---|---|---|---|
| Energy (Kcal) | Male | 1683.0 (704.4-3403) | 0.551 | 1779.0 (693.9-3315) | 0.172 |
| | Female | 1481.0 (733.3-2860) | | 1540.0 (812-2527) | |
| Carbohydrate (g) | Male | 281.6 (119.2-726.1) | 0.291 | 296.7 (91.9-2331) | 0.058 |
| | Female | 232.0 (112.2-459.9) | | 244.3 (108.6-406.7) | |
| Protein (g) | Male | 33.64 (14.5-76.2) | 0.533 | 45.8 (8.1-89.3) | 0.381 |
| | Female | 34.2 (18.0-81.1) | | 38.9 (11.6-111.3) | |
| Fats (g) | Male | 33.7 (14.6-88.8) | 0.184 | 44.9 (20.3-104.2) | 0.459 |
| | Female | 42.9 (10.6-137.9) | | 43.9 (18.7-79.3) | |
| Fibre (g) | Male | 20.8 (7.2-49.8) | 0.763 | 26.0 (7.6-49.9) | 0.262 |
| | Female | 25.5 (9.9-43.8) | | 19.5 (9.4-45.5) | |
| Folate (µg) | Male | 267.4 (79.5-928.8) | 0.995 | 343.7 (4.2-916.5) | **0.018** |
| | Female | 298.4 (96.8-710.1) | | 214.6 (61.8-475.1) | |
| Iron (mg) | Male | 9.93 (2.2-18.5) | 0.733 | 11.2 (3.7-23.7) | 0.100 |
| | Female | 8.9 (4.7-27.3) | | 9.3 (4.2-18.6) | |
| Selenium (µg) | Male | 46.5 (16.9-128.8) | 0.683 | 77.0 (28.4-176.8) | 0.240 |
| | Female | 43.3 (29.0-197.3) | | 66.7 (13.3-231.1) | |
| Vitamin A (µg) | Male | 423.1 (6.0-1852) | 0.587 | 437.5 (12.04-3778.0) | 0.155 |
| | Female | 282.1 (7.81-2153) | | 281.6 (17.71-1575) | |
| Vitamin $B_{12}$ (µg) | Male | 0.6 (0.0-4.1) | 0.899 | 2.0 (0.0-11.5) | 0.296 |
| | Female | 0.8 (0.0-3.0) | | 1.0 (0.14-8.7) | |
| Vitamin C (mg) | Male | 91.9 (35.4-432.4) | 0.743 | 104.4 (23.4-247.6) | 0.209 |
| | Female | 75.3 (33.00-301) | | 64.3 (32.2-223.7) | |
| Vitamin E (mg) | Male | 5.0 (2.3-8.9) | 0.952 | 6.2 (1.7-13.5) | 0.237 |
| | Female | 4.8 (1.5-22.14) | | 5.650 (2.05-8.9) | |
| Vitamin K (µg) | Male | 44.5 (9.1-109.5) | 0.743 | 38.2 (4.3-211.3) | 0.396 |
| | Female | 22.0 (5.8-137.2) | | 28.5 (2.3-119.1) | |
| Zinc (mg) | Male | 5.6 (2.4-7.8) | 0.606 | 7.4 (3.5-12.5) | 0.155 |
| | Female | 5.9 (2.5-9.4) | | 5.3 (1.7-14.14) | |

Data were compared in cases and controls between males and females using a Mann-Whitney test. P-value <0.05 are indicated in bold text.

**S6 Table: Comparison of energy and nutrient intake of Cohort 1 study participants between those with different household income.**

| Nutrients | Group | Household monthly income<br><Gh¢50<br>BU cases N=6<br>Controls N=4<br>Median (range) | Gh¢50-200<br>BU cases N=10<br>Controls N=15<br>Median (range) | Gh¢201-800<br>BU cases N=24<br>Controls N=21<br>Median (range) | P-value |
|---|---|---|---|---|---|
| Energy (Kcal) | BU cases | 1165 (914-1876) | 1629 (926-2734) | 1525 (704-3403) | 0.690 |
| | Controls | 1401 (694-3315) | 1732 (812-3296) | 1553 (723-2828) | 0.763 |
| Carbohydrate (g) | BU cases | 209.0 (136-329) | 276.0 (119-444) | 249.0 (112-726) | 0.714 |
| | Controls | 209.0 (91.9-558) | 290.0 (109-460) | 253.0 (113-2331) | 0.712 |
| Protein (g) | BU cases | 28.1 (26.2-38.4) | 39.4 (23.2-61.8) | 34.5 (14.5-81.1) | 0.405 |
| | Controls | 32.6 (8.15-73.4) | 41.2 (24.7-111) | 45.1 (11.6-75.8) | 0.684 |
| Fats (g) | BU cases | 45.3 (14.6-51.7) | 37.6 (22.9-138) | 36.0 (10.6-94.6) | 0.900 |
| | Controls | 51.8 (33.8-98.1) | 42.8 (29.1-104) | 44.9 (18.7-84.0) | 0.492 |
| Fibre (g) | BU cases | 21.3 (10.0-33.6) | 24.2 (9.95-41.1) | 24.4 (7.17-49.8) | 0.983 |
| | Controls | 18.3 (9.1-49.4) | 24.8 (10.6-49.9) | 21.7 (7.59-49.4) | 0.875 |
| Folate (µg) | BU cases | 263 (173-382) | 231 (90.5-518) | 269 (79.5-929) | 0.756 |
| | Controls | 319 (253-512) | 327 (4.2-604) | 238 (61.8-916) | 0.200 |
| Iron (mg) | BU cases | 7.2 (5.5-11.4) | 10.2 (3.9-18.5) | 9.1 (2.2-27.3) | 0.633 |
| | Controls | 6.8 (4.5-16.6) | 12.0 (4.5-19.0) | 8.9 (3.7-23.7) | 0.197 |
| Selenium (µg) | BU cases | 42.6 (29.0-43.0) | 50.2 (30.7-129) | 44.9 (16.9-197) | 0.126 |
| | Controls | 76.1 (36.8-177) | 79.2 (29.8-231) | 65.7 (13.3-142) | 0.284 |
| Vitamin A (µg) | BU cases | 554.0 (282-1307) | 614.0 (7.82-2153) | 145.0 (6.00-2153) | 0.168 |
| | Controls | 427.0 (31.9-1631) | 392.0 (18.1-2174) | 278 (12.0-3778) | 0.785 |
| Vitamin $B_{12}$ (µg) | BU cases | 0.815 (0.100-2.99) | 0.815 (0.0-2.06) | 0.520 (0.00-4.14) | 0.975 |
| | Controls | 1.98 (0.32-2.66) | 1.86 (0.54-9.01) | 0.94 (0.00-11.5) | 0.652 |
| Vitamin C (mg) | BU cases | 130 (60.0-186) | 87.0 (39.1-301) | 97.9 (33.0-432) | 0.854 |
| | Controls | 115 (66.1-157) | 62.0 (23.4-248) | 105 (33.9-224) | 0.068 |
| Vitamin E (mg) | BU cases | 6.62 (2.76-7.27) | 4.87 (2.05-22.1) | 4.58 (1.50-13.1) | 0.662 |
| | Controls | 8.1 (6.03-13.6) | 6.0 (1.69-12.0) | 5.4 (2.05-13.1) | 0.221 |
| Vitamin K (µg) | BU cases | 63.1 (8.29-106) | 34.9 (7.97-137) | 33.5 (5.83-137) | 0.440 |
| | Controls | 92.7 (40.7-153) | 30.0 (4.27-195) | 27.5 (2.72-211) | 0.089 |
| Zinc (mg) | BU cases | 4.95 (2.55-6.16) | 5.70 (3.31-7.30) | 5.86 (2.36-9.38) | 0.298 |
| | Controls | 5.16 (3.66-12.0) | 7.21 (3.74-14.1) | 6.24 (1.70-11.4) | 0.445 |

Data were compared in cases and controls between those with different levels of household monthly income, using the Kruskal-Wallis test and Dunn's post-hoc test for multiple comparisons.
